# A multicentre study to predict COVID-19 outbreaks in long-term care homes using wastewater surveillance and environmental surface sampling for SARS-CoV-2

**DOI:** 10.1101/2025.10.20.25337094

**Authors:** Jason A. Moggridge, Kritisha Acharya, Derek R. MacFadden, Alex Wong, Rees Kassen, Caroline Nott, Gustavo Ybazeta, David S. Guttman, Lucas Castellani, Michael Fralick

**Author notes:** Correspondence: Michael Fralick, MD, PhD, SM, FRCPC, Sinai Health System, 60 Murray Street, M5T 3L9. **Study concept and design:** All authors. **Analysis/interpretation of data:** All authors. **Drafting of the manuscript:** Moggridge JA, Fralick M, Acharya K. **Critical revision of the manuscript:** All authors. **Statistical analysis:** Moggridge JA.

## Abstract

**Background:** Floor swabs can be an effective environmental sampling method for proactive SARS-CoV-2 surveillance in congregate settings like long-term care homes (LTCHs). Concurrent assessment of additional variables such as wastewater surveillance data and weather data have the potential to improve the predictive performance of this approach.

**Methods:** We analyzed existing data from 5,095 floor swabs collected between August 2021 and January 2023 from 10 LTCHs across three cities in Ontario, Canada: Ottawa, Toronto, and Sault Ste. Marie. Floors were swabbed weekly at each LTCH. Swabs were analyzed using RT-qPCR. Wastewater data was obtained from the Ontario Wastewater Surveillance Consortium’s repository; we included one treatment plant for each city. Weather data was sourced from Environment Canada, with one station selected from each city. Logistic regression, LASSO-penalized logistic regression, Random Forest, and XGBoost were used for COVID-19 outbreak predictions using different subsets of predictors with leave-one-LTCH-out cross-validation. SHAP values were computed for model explainability. Our outcome of interest was a COVID-19 outbreak within an LTCH.

**Results:** Over the study period, 25 COVID-19 outbreaks occurred in the participating LTCHs, with a median duration of 30 days and a median of 39 cases per outbreak (range 2 to 196). LASSO generally out-performed logistic regression, Random Forest, and XGBoost. The two variables with the highest SHAP values were log transformed 7-day mean wastewater and log viral copies from floor swabs.

**Conclusions:** Incorporating wastewater data and weather data enhanced the ability of floor swab results to predict an outbreak of COVID-19 in an LTCH. Future studies are needed to evaluate how well the model performs when implemented into practice.

## INTRODUCTION

Residents of long term care homes (LTCHs) were disproportionately affected by COVID-19, and experienced high rates of morbidity and mortality.^1,2^ At the beginning of the pandemic, the mortality rate from COVID-19 outbreaks in Canadian LTCHs exceeded 30%, with LTC residents comprising 81% of all COVID-19 deaths in Canada.^3,4^ The outbreaks also had other adverse effects. For example, there was limited access to outpatient services or visiting healthcare professionals for residents, restrictions on community groups and day programs, and social isolation. To mitigate the impact of COVID-19 outbreaks on LTCHs, consistent and proactive disease surveillance is crucial in predicting and preventing future outbreaks. While active clinical testing is a commonly used approach, it is costly and hard to sustain.^5^ Passive surveillance, like environmental detection, offers a non-invasive option for congregate settings.

Our team has validated a method of built environment sampling for SARS-CoV-2 in which swab samples are taken from floors and processed using reverse transcription quantitative polymerase chain reaction (RT-qPCR).^6^ Our largest study was a 14-month, multicentre prospective study at 10 LTCHs in Ontario, which yielded three main findings.^7^ First, the percentage of floor swabs detecting SARS-CoV-2 can effectively rule out an active outbreak. Second, the percentage of floor swabs positive for SARS-CoV-2 rises days, and sometimes weeks, before an outbreak is identified. Lastly, floor swabs provide spatial resolution to identify the areas in the LTCH where COVID-19 cases occur. Our data suggest that floor swabs can serve as a proactive surveillance method. However, this analytic approach is primarily descriptive, and while viral load is a strong predictor of COVID-19 burden, each LTCH also has specific factors that can either contribute to or hinder the transmission of SARS-CoV-2. In addition, prior studies have shown that weather (e.g., temperature, wind speed) are associated with the spread of COVID-19.^8,9^ For example, a recent review article highlighted that multiple studies have shown a negative correlation between ambient temperature and number of COVID-19 cases.^9^ The objective of our current study was to include additional predictors such as local wastewater data and weather to identify whether these variables improved model performance.

## METHODS

### Study Design

To enhance the predictive accuracy of detecting COVID-19 outbreaks in LTCHs, we utilized our existing floor swab RT-qPCR results with supplementary data from regional wastewater monitoring and weather stations. Our study was reviewed by the research ethics board at the University of Ottawa and received a waiver because the samples collected are not human (as discussed in Article 2.1 of the Tri-Council Policy Statement: Ethical Conduct for Research Involving Humans [TCPS 2]), and any demographic data are either: (1) publicly available through a mechanism set out by legislation or regulation and that is protected by law; or (2) they are in the public domain and the individuals to whom the information refers have no reasonable expectation of privacy (see Article 2.2 of the TCPS 2).

### Data Sources

Between August 2021 and January 2023, our team collected and assayed by RT-qPCR 5,095 floor swabs from 10 LTCHs across three cities in Ontario, Canada: Ottawa, Toronto, and Sault Ste. Marie. Floors were swabbed weekly in the same interior locations during each visit. Each LTCH was blinded to the results. All other methods related to floor swab collection, RNA extraction, and RT-qPCR testing have been described previously.^7^ Results for each collection date were aggregated as the rate of positive tests (positivity) and the geometric mean number of SARS-CoV-2 RNA copies detected plus one. The pseudocount of one copy was added to each value of viral copies to enable logarithmic transformation of these data (as the logarithm of zero is undefined) for summary statistics, visualization, and modelling.

Confirmed COVID-19 outbreaks in LTCHs were defined as “two or more lab-confirmed COVID-19 cases in residents, staff or other visitors in a home, with an epidemiological link, within a 14-day period, where at least one case could have reasonably acquired their infection in the LTC home” as per provincial guidance in Ontario set before the start of the sample collection period.^10^ The start date, end date, and number of COVID-19 cases for residents and staff were recorded for each outbreak. Over the study period, 25 outbreaks occurred, with a median duration of 30 days and a median of 39 cases per outbreak (range 2 to 196).

We accessed historical SARS-CoV-2 wastewater data from the areas surrounding the participating LTCHs. The wastewater sampling frequency (both over time and among sites) ranged from biweekly to daily, with each sampling instance reporting wastewater viral concentrations. All wastewater data was obtained from the Ontario Wastewater Surveillance Consortium’s repository.^11^ We selected results from the relevant treatment plant (TP) sites for each city: “Ottawa WWTP” (ROPEC TP facility) in Ottawa, “AB” (Ashbridges Bay TP) in Toronto, and “TPSSM” in Sault Ste. Marie. All wastewater results were from qPCR assays of solid fraction samples. The mean concentration of N1 and N2 gene targets was normalized against the concentration of pepper mild mottle virus (PMMoV) to compute daily relative SARS-CoV-2 proportions for each city. These daily relative proportions were used to calculate 7-day endpoint averages (with missing values ignored) for modelling. We used the last-observation-carried-forward method to replace missing values for a small number of dates with an undefined rolling average.

Weather data were downloaded from Environment Canada.^12^ We selected a single, representative weather station for each city. Station names and IDs for these were OTTAWA CDA RCS (#30578), TORONTO INTL A (#51459), and SAULT STE MARIE A (#50092). We collected data for the following variables (units): daily mean temperature (°C), total precipitation (mm), average relative humidity (%), speed (kph) and direction (in degrees) of the maximum wind gust, and barometric pressure (mbar). We computed each variable’s 7-day endpoint rolling averages and 7-day spreads (i.e., max. value - min. value); however, spread was not computed for wind direction. These data were used as predictors for modelling COVID-19 outbreaks in LTCHs.

### Study Outcome and Covariates

The primary outcome variable was the presence of a COVID-19 outbreak within an LTCH as determined by local public health guidelines. Model covariates include floor swab results (i.e., test positivity and viral copies recovered), viral detection in wastewater, and weather data.

### Statistical Analysis

All statistical analyses were performed using the R programming language (v4.4.1). We computed descriptive statistics, as well as Pearson correlation coefficients between each covariate, and between covariates and COVID-19 outbreaks. Confidence intervals for continuous variables were calculated using the Wald method, and the Agresti-Coull method was used for confidence intervals of binary variables. Finally, we evaluated various supervised machine learning classifiers for outbreak discrimination, with outbreak status modelled as a binary outcome and concurrent results from LTCH floor swabs, regional wastewater testing, and weather records used as predictors.

### Supervised Learning Methods

We evaluated classifiers created using several different supervised learning methods, including logistic regression using the *glm* package, LASSO regression using the *glmnet* package (abbreviated as LR in this work), Random Forest using the *ranger* package (RF), and boosted trees with the *xgboost* package (XGB). Classifiers were evaluated by leave-one-LTCH-out cross-validation (k=10 LTCHs), where each fold corresponded to the set of observations from a single LTCH, and each of the ten iterations had a different set of observations from a single LTCH held out for testing.

For each iteration in cross-validation, we applied a data pre-processing in which categorical predictors were converted to dummy variables, centring and scaling was performed for all numeric predictors, and any zero-variance variables were dropped from the dataset. To prevent data leakage, learned pre-processor settings (specifically, means and standard deviations for centring and scaling) were determined in each iteration from the training examples only.

Optimal hyperparameters for each method were determined in cross-validation. For LR, we tuned the LASSO regularization penalty λ; for RF, we searched for the optimal combination of *mtry*, the number of predictors sampled at each split and *min_n*, the minimum number of observations required per node for further splitting; for XGB, we tuned *mtry, min_n*, tree-depth, learning-rate (shrinkage), loss-reduction required for further splits, sample-size used for fitting, and the number of iterations with improvement before stopping. For LR and RF, we used a grid-search with 250 and 25 different hyperparameter settings, respectively. For XGB, we used a Latin hypercube search with 100 iterations, due to the larger number of hyperparameters requiring tuning.

To assess the performance of our predictive model, we chose key metrics such as the area under the receiver operating curve (AUROC), accuracy, and Brier score. Model calibration was assessed by plotting the predicted probability of an outbreak against the observed probability of an outbreak. Receiver operator curves were plotted by varying the alarm threshold for the predicted probability from 0.1% to 99.9%. Additionally, we have provided SHAP (SHapley Additive exPlanations) values to estimate the relative importance of each included variable to aid in model interpretability.^13,14^ These were calculated using the *kernelshap* package.

### RESULTS

We collected and assayed 5,095 floor samples from 10 LTCHs across Ottawa, Toronto, and Sault Ste. Marie in Ontario, Canada between August 2021 and January 2023 (Table 1). The number of samples collected per facility ranged from 148 to 1,157. This variability is attributed to the size of the LTCH and the duration of the monitoring period. The monitoring periods at different LTCHs ranged from 112 days (with 15 sample collection visits) to 428 days (with 56 visits). Overall, SARS-CoV-2 RNA was detected by RT-qPCR in 1,808 of 5,095 samples (35.5%, 95% CI: 34.0, 37.0%), and the geometric mean number of genomic copies recovered per assay (plus one) was 2.57 (95% CI: 2.5, 2.7 copies). Aggregate SARS-CoV-2 RNA detection rates varied among LTCHs from 10.8% (CI: 6.7, 17.0%) for the LTCH with the least test positivity to 57.5% (CI: 54.0, 61.0%) for the LTCH with the greatest positivity. The aggregate geometric mean number of viral copies (plus one) detected in all samples varied from 1.14 (CI: 1.07, 1.2) to 6.3 (CI: 5.5, 7.1) among LTCHs. Characteristics of the LTCH buildings have been previously described.^7^

**Table 1:** Summary statistics for floor swabs at each long-term care home.

| Site | # of floor swabs | # of swabs positive for SC2 | Start date | End date | Visits | Proportion of swabs positive for SC2 | SC2 Viral Copies |
| --- | --- | --- | --- | --- | --- | --- | --- |
| **Overall** |  |  |  |  |  |  |  |
|  | 5,095 | 1,808 | 2021-08-30 | 2023-01-06 | 163 | 0.355<br>(0.342, 0.368) | 2.57<br>(2.46, 2.68) |
| **By LTCH** |  |  |  |  |  |  |  |
| LTC-01 | 633 | 131 | 2021-09-07 | 2022-05-26 | 38 | 0.207<br>(0.177, 0.24) | 1.47<br>(1.37, 1.58) |
| LTC-02 | 629 | 154 | 2021-08-31 | 2022-05-26 | 37 | 0.245<br>(0.213, 0.28) | 1.64<br>(1.49, 1.8) |
| LTC-03 | 570 | 197 | 2021-08-30 | 2022-05-30 | 38 | 0.346<br>(0.308, 0.386) | 2.1<br>(1.88, 2.33) |
| LTC-04 | 1157 | 424 | 2021-11-04 | 2023-01-06 | 56 | 0.366<br>(0.339, 0.395) | 2.63<br>(2.4, 2.88) |
| LTC-05 | 1010 | 581 | 2022-01-25 | 2023-01-06 | 43 | 0.575<br>(0.545, 0.605) | 6.28<br>(5.54, 7.11) |
| LTC-06 | 265 | 101 | 2021-11-03 | 2022-03-16 | 19 | 0.381<br>(0.325, 0.441) | 3.55<br>(2.82, 4.48) |
| LTC-07 | 148 | 16 | 2021-10-29 | 2022-02-18 | 15 | 0.108<br>(0.0667, 0.169) | 1.14<br>(1.07, 1.21) |
| LTC-08 | 260 | 41 | 2021-11-04 | 2022-03-14 | 20 | 0.158<br>(0.118, 0.207) | 1.25<br>(1.17, 1.34) |
| LTC-09 | 190 | 99 | 2021-11-05 | 2022-03-07 | 19 | 0.521<br>(0.45, 0.591) | 4.89<br>(3.71, 6.46) |
| LTC-10 | 233 | 64 | 2021-11-01 | 2022-03-16 | 31 | 0.275<br>(0.221, 0.335) | 1.81<br>(1.55, 2.11) |

Legend: SC2 = SARS-CoV-2, LTCH = long-term care home. Numbers in brackets represent 95% confidence interval.

### Description of wastewater SARS-CoV-2 monitoring and weather records

Daily regional wastewater signals relative to PMMoV were smoothed for modelling using a 7-day endpoint rolling average. Daily wastewater detection of SARS-CoV-2 was greater at Sault Ste. Marie (median: 0.0038 copies / PMMoV; IQR: 0.0015, 0.0077) than Toronto (3.1 × 10^-4^; IQR: 6 × 10^-5^, 8.5 × 10^-4^) or Ottawa (1.3 × 10^-4^, IQR: 6.12 × 10^-5^, 2.8 × 10^-4^), though the sampling periods were substantially different for each city (Table 2).

**Table 2.** City-level characteristics relevant to multiple long-term care homes, including SARS-CoV-2 concentrations in wastewater and weather variables.

|  | **Ottawa** | **Sault Ste. Marie** | **Toronto** |
| --- | --- | --- | --- |
| Start date | 2021-10-29 | 2021-11-04 | 2021-08-30 |
| End date | 2022-03-16 | 2023-01-06 | 2022-05-30 |
| Precipitation (mm) | 0 (0, 1.95) | 0.2 (0, 3.48) | 0 (0, 1.85) |
| Mean temp (°C) | -5.3 (-12.2, 0.2) | 2.9 (-4.2, 13.2) | 5.9 (-0.3, 12.8) |
| Max. temp (°C) | -0.6 (-6.1, 4.5) | 7 (-0.4, 19.9) | 9.4 (2.8, 17.8) |
| Min. temp (°C) | -9.4 (-17.9, -3.7) | -1.3 (-7.3, 6.2) | 1.1 (-3.8, 8.1) |
| Max wind speed (km/h) | 34 (28, 44) | 39 (28, 50) | 41 (33, 52) |
| Mean pressure (kPa) | 100.73 (100.12, 101.42) | 99.19 (98.64, 99.73) | 99.52 (99.0, 100.08) |
| Relative humidity (%) | 73 (62, 84) | 82 (69, 94) | 71 (60, 83) |
| Wastewater SC2:PMMoV | $1.3 \times 10^{-4}$<br>($6.12 \times 10^{-5}$, $2.8 \times 10^{-4}$) | $3.8 \times 10^{-3}$<br>($1.5 \times 10^{-3}$, $7.7 \times 10^{-3}$) | $3.1 \times 10^{-4}$<br>($6 \times 10^{-5}$, $8.5 \times 10^{-4}$) |

Legend: SC2 = SARS-CoV-2, PMMoV = pepper mild mottle virus. Results are aggregates of per-day statistics (e.g., daily mean temperature, etc.) expressed as median (IQR), except for dates.

We collected weather data from three Environment Canada stations near the 10 LTCHs (Table 2). We used temperature, precipitation, and wind statistics from the daily records and calculated daily statistics for barometric pressure and relative humidity using hourly observations. Toronto had the highest average daily temperatures (min = 1.2°C, mean = 5.6°C, and max = 10.1°C), followed by Sault Ste. Marie (min = -1.7°C, mean = 3.6°C, max = 9.0°C) and Ottawa (min = - 10.8°C, mean = -5.8°C, max = -0.8°C). Toronto was also the windiest, with an average daily maximum wind gust speed of 43.9 km/h, followed by Sault Ste. Marie (42.1 km/h) and Ottawa (37.5 km/h). Sault Ste. Marie had the largest average daily precipitation with 2.6 mm, followed by Toronto (2.4 mm) and Ottawa (1.8 mm). Ottawa had the highest average hourly barometric pressure (100.7 kPa), followed by Toronto (99.5 kPa) and Sault Ste. Marie (99.2 kPa). Sault Ste. Marie had the greatest average hourly relative humidity (79.4%), followed by Ottawa (72.8%) and Toronto (71.2%).

### Pairwise Correlations

We computed Pearson correlations for all pairs of variables used in modelling (Suppl. Figure S2). Significant correlations were found between floor swab positivity and viral copies (r = 0.91), floor swab positivity and wastewater signal (7-day rolling endpoint average; r = 0.45), and floor swab positivity and daily temperature range (7-day rolling endpoint average; r = 0.3). Floor swab viral copies were also significantly correlated with wastewater signal (r = 0.52), as well as temperature range (r = 0.31). Wastewater signal was positively correlated with daily temperature range (r = 0.23), daily wind speed range (7-day endpoint average; r = 0.15), but negatively correlated with barometric pressure (r = -0.51).

### ML Model performance

We evaluated outbreak discrimination by single logistic regression with three different predictor variables to establish benchmarks for machine-learning models (Figure 1). Floor swab qPCR results provided the best predictors of outbreaks in leave-one-LTCH-out cross-validation: floor swab viral copies had an AUROC of 0.82 ± 0.05, accuracy of 0.72 ± 0.05, and Brier score of 0.19 ± 0.03, while floor swab test positivity had similar performance (AUROC: 0.81 ± 0.04; accuracy: 0.73 ± 0.04; Brier: 0.19 ± 0.03). Regional wastewater (as 7-day rolling endpoint average) had comparable AUROC (0.82 ± 0.05) but worse accuracy (0.62 ± 0.05) and Brier score (0.23 ± 0.03) than the floor-swab–derived measures.

**Figure 1.**
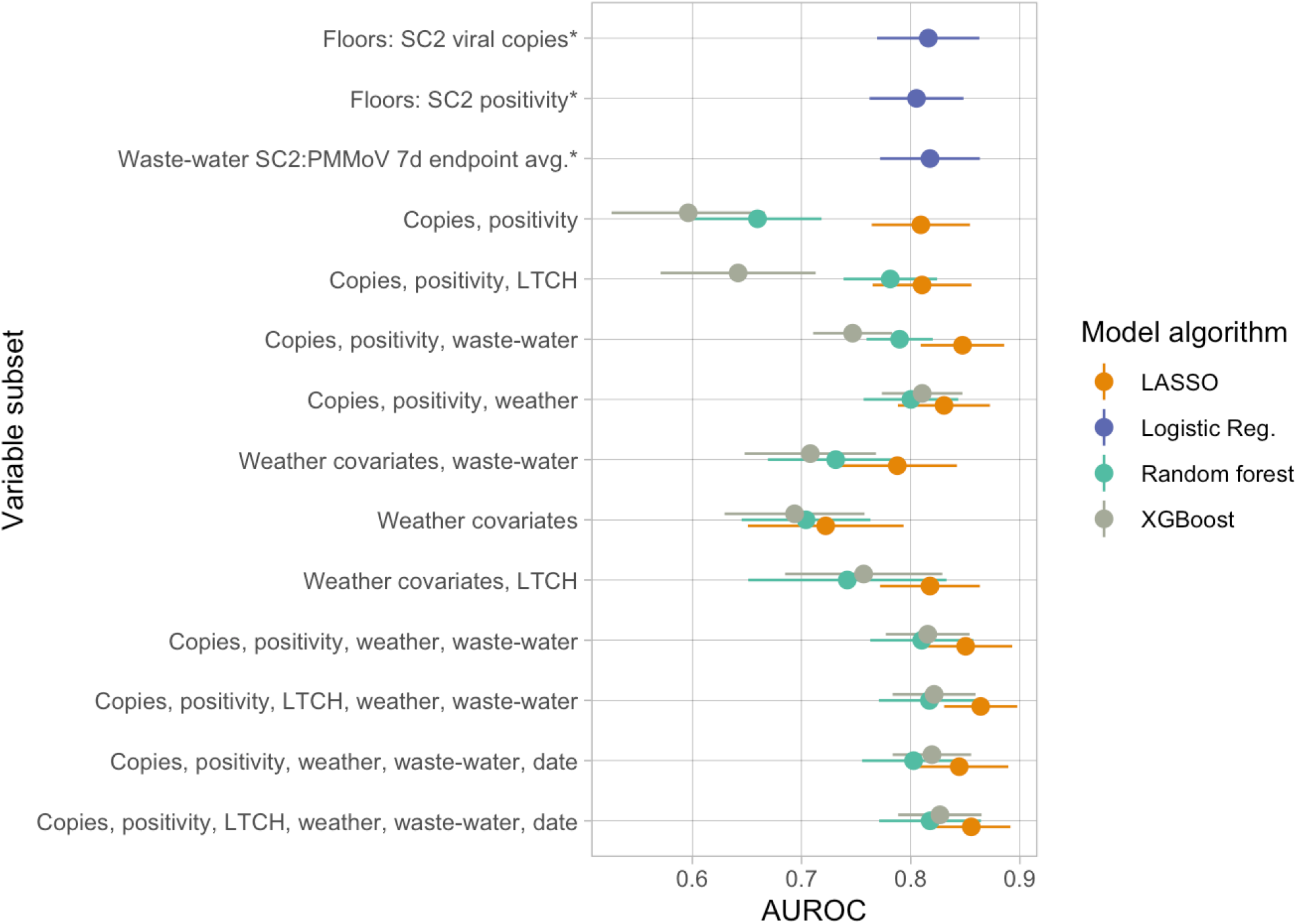
AUROC results for different combinations of machine learning methods and subsets of predictors, evaluated by leave-one-LTCH-out cross-validation (k=10 sites). SC2 = SARS-CoV-2, PMMoV = pepper mild mottle virus, 7d = 7-day, LTCH = long-term care home, AUROC = area under the receiver operating curve

We evaluated more complex predictive models using three different machine learning methods (LASSO logistic regression, Random Forest, and XGBoost) and various subsets of available predictors, including aggregate floor swab test results with and without dummy variables for different LTCH sites, regional wastewater signals, and weather variables (Figure 1; Suppl. Table S2). A LASSO regression using only floor swab positivity and viral copies with dummy variables for different LTCHs performed similarly to the logistic regression with either predictor alone, with AUROC of 0.81 ± 0.05. Random Forest and XGBoost with the same predictors performed slightly worse (AUROC 0.79 ± 0.04 and 0.80 ± 0.04, respectively). Including wastewater as an additional predictor significantly improved the performance of LASSO (0.85 ± 0.04) and XGBoost (0.87 ± 0.03) models. Maximal performance in terms of AUROC occurred when swab results, weather, and wastewater variables were used with LTCH dummy variables, with LASSO performing slightly better than the other two methods (LASSO: 0.88 ± 0.03; RF: 0.86 ± 0.03; XGB: 0.87 ± 0.03). Overall, including additional weather and wastewater predictors yielded a slight improvement in outbreak prediction compared to simple logistic regression with either floor swab test positivity or viral copies as a single predictor (Suppl. Table S2).

We selected the LASSO model that maximized AUROC in cross-validation as the “best” model and performed analysis of SHAP values. For this model, the mean absolute SHAP values were (from largest to smallest) wastewater 7-day rolling average (0.165), LTCH dummy variables (0.134), floor swab viral copies (0.127), floor swab positivity (0.048), wind direction (0.040), wind speed (0.023), mean temperature (0.020), relative humidity (0.010), mean barometric pressure (0.007), and finally weather variables that appear to be low utility predictors having the same 0.003 mean SHAP value: wind speed range, temperature range, precipitation, relative humidity range, and barometric pressure range (Figure 2).

**Figure 2.**
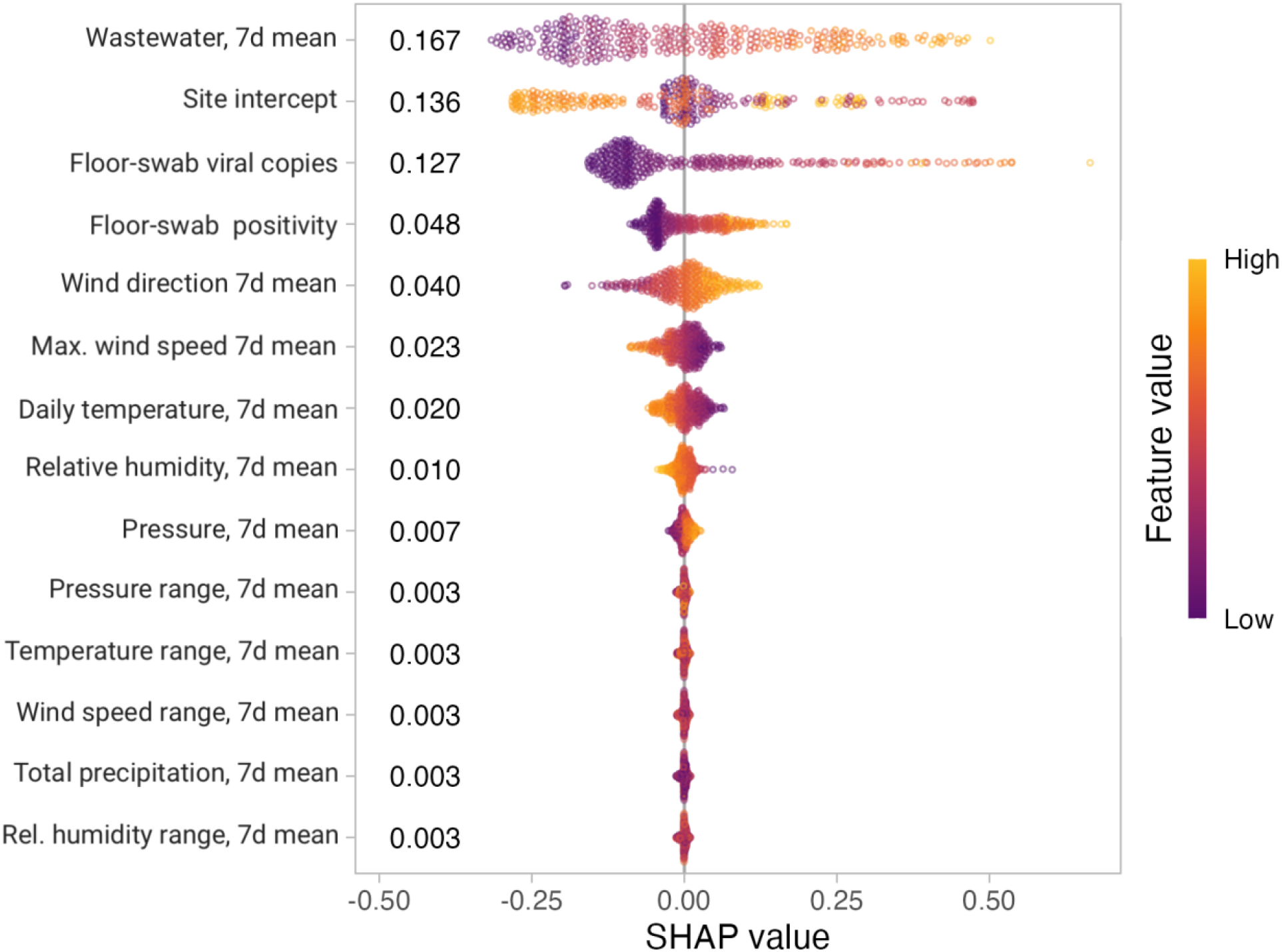
SHAP values for each predictor of outbreak status from the selected LASSO model.

## DISCUSSION

In this data analysis from a multicentre prospective study across 10 LTCs,^7^ the combination of floor swabs, weather data, and wastewater data could accurately predict a COVID-19 outbreak. The predictions were generated with three of the most commonly used supervised machine learning techniques. The results highlight how the combination of advanced analytic approaches and unique data sources can be leveraged to predict outbreaks of COVID-19.

Over the course of the pandemic, wastewater has received significant attention and funding for its role in estimating disease burden in a given region. Its primary use case has been for surveillance, as opposed to real-time prediction and prevention, for three main reasons. First, in general there is a lag of days (and sometimes weeks) between collection of wastewater samples and availability of results. Second, most often wastewater results are reported on a regional level, and it is rare for results to be available for an individual LTCH or hospital. As a result, wastewater generally lacks spatial resolution; although we acknowledge it is technically possible for an individual building to have its own dedicated wastewater surveillance. In contrast, floor swabbing has lower resource demands and can provide spatial resolution to the level of a single room within a building. A recent pilot demonstrated that floor swab results can be available within 48 hours of a building being swabbed.^15^ However, a wider implementation study of floor sampling for SARS-CoV-2 outbreak prediction and prevention has not yet been conducted. The results of this current study suggest that the addition of weather and wastewater data can improve the predictive performance of floor swabs.

Our study has four main strengths. First, it was multicentre and conducted in both urban and community settings. Second, it incorporated diverse datasets and provided estimates for the relative importance of each of the included variables. Third, it applied supervised machine learning techniques which are designed for prediction. Fourth, it represents one of the few studies that directly compared wastewater sampling to environmental surface sampling (i.e., floor swabbing).

Our study also has important limitations. First, while our model was built with both prospective data (e.g., floor sampling) and retrospective data (e.g., wastewater data), a future prospective study is needed to identify whether acting on these predictions can mitigate the size and scope of an outbreak. Second, our study included both urban and community settings, but it is unknown how our results might generalize to rural settings or regions with less climate variability. Third, our study was restricted to LTCHs, and thus it is unknown how results might generalize to other congregate settings such as retirement homes or acute care hospitals. Fourth, we did not have access to data on other respiratory viruses such as influenza or respiratory syncytial virus (RSV). These viruses commonly cause outbreaks and have high mortality rates in LTCHs, which underscores the importance of surveillance methods that consider multiple viruses in addition to SARS-CoV-2. Fifth, we did not conduct a cost-effectiveness study to help assess whether this approach is affordable or sustainable for a LTCH.

In closing, the results of our study suggest that incorporating wastewater data and weather data enhances the ability of floor swab results to predict a COVID-19 outbreak in an LTCH. Future work is needed to evaluate whether acting on such results can mitigate the size and scope of COVID-19 outbreaks in LTCHs.

## Data Availability

Results of environmental surveillance by swabbing location cannot be shared due to data and privacy agreements. All wastewater data were obtained from the Ontario Wastewater Surveillance Consortium's publicly available repository at https://github.com/OntarioWastewaterSurveillanceConsortium/sars-cov-2-data. Weather data were downloaded from Environment Canada's publicly available Historical Climate Data database at https://climate.weather.gc.ca/.

https://climate.weather.gc.ca/

