## Supplementary Materials for "A multicentre study to predict COVID-19 outbreaks in long-term care homes using wastewater surveillance and environmental surface sampling for SARS-CoV-2"

### Supplemental Materials

**Supplemental Table S1.** Summary statistics for swabs by current long-term care home (LTCH) outbreak status.

| Outbreak | Total swabs (n) | SC2 Positives (n) | Duration (days) | Visits (n) | SC2 Positivity (mean, 95% CI) | Copies (mean, 95% CI) |
|---|---|---|---|---|---|---|
| No | 3,170 | 729 | 494 | 130 | 0.23 (0.216, 0.245) | 1.67 (1.61, 1.74) |
| Yes | 1,925 | 1079 | 402 | 78 | 0.561 (0.538, 0.583) | 5.22 (4.78, 5.69) |

Legend: SC2 = SARS-CoV-2, 95% CI = 95% Confidence Interval.

**Supplemental Table S2.** Performance metrics (accuracy, Brier score, and AUROC) summarized for each combination of method and predictor(s) subset tested in cross-validation. Values are expressed as mean ± standard error.

| Predictors | LASSO | Random Forest | XGBoost |
|---|---|---|---|
| **Accuracy** | | | |
| Floors: SC2 viral copies* | 0.72 ± 0.05 | NA | NA |
| Floors: SC2 positivity* | 0.73 ± 0.04 | NA | NA |
| Waste-water SC2:PMMoV 7-day endpoint average* | 0.62 ± 0.05 | NA | NA |
| Copies, positivity | 0.72 ± 0.05 | 0.70 ± 0.05 | 0.72 ± 0.05 |
| Copies, positivity, LTCH | 0.72 ± 0.05 | 0.72 ± 0.05 | 0.72 ± 0.05 |
| Copies, positivity, wastewater | 0.71 ± 0.05 | 0.70 ± 0.04 | 0.72 ± 0.04 |
| Copies, positivity, weather | 0.72 ± 0.04 | 0.71 ± 0.04 | 0.71 ± 0.04 |
| Weather covariates, wastewater | 0.64 ± 0.05 | 0.66 ± 0.06 | 0.66 ± 0.06 |
| Weather covariates | 0.63 ± 0.05 | 0.67 ± 0.05 | 0.69 ± 0.04 |
| Weather covariates, LTCH | 0.60 ± 0.05 | 0.65 ± 0.07 | 0.67 ± 0.05 |
| Copies, positivity, weather, wastewater | 0.70 ± 0.04 | 0.73 ± 0.04 | 0.76 ± 0.03 |
| Copies, positivity, LTCH, weather, wastewater | 0.70 ± 0.05 | 0.73 ± 0.04 | 0.77 ± 0.03 |
| Copies, positivity, weather, wastewater, date | 0.69 ± 0.05 | 0.74 ± 0.04 | 0.73 ± 0.04 |
| Copies, positivity, LTCH, weather, wastewater, date | 0.70 ± 0.05 | 0.73 ± 0.04 | 0.73 ± 0.04 |
| **Brier score (lower scores indicate better performance)** | | | |
| Floors: SC2 viral copies* | 0.19 ± 0.03 | NA | NA |
| Floors: SC2 positivity* | 0.19 ± 0.03 | NA | NA |
| Wastewater SC2:PMMoV 7-day endpoint average* | 0.23 ± 0.03 | NA | NA |
| Copies, positivity | 0.19 ± 0.03 | 0.21 ± 0.03 | 0.20 ± 0.015 |
| Copies, positivity, LTCH | 0.19 ± 0.04 | 0.19 ± 0.03 | 0.20 ± 0.015 |
| Copies, positivity, wastewater | 0.20 ± 0.03 | 0.18 ± 0.03 | 0.19 ± 0.03 |
| Copies, positivity, weather | 0.19 ± 0.03 | 0.18 ± 0.02 | 0.18 ± 0.016 |

| | | | |
|---|---|---|---|
| Weather covariates, wastewater | 0.23 ± 0.016 | 0.20 ± 0.02 | 0.20 ± 0.016 |
| Weather covariates | 0.23 ± 0.016 | 0.21 ± 0.02 | 0.21 ± 0.014 |
| Weather covariates, LTCH | 0.27 ± 0.05 | 0.22 ± 0.04 | 0.20 ± 0.02 |
| Copies, positivity, weather, wastewater | 0.18 ± 0.03 | 0.17 ± 0.02 | 0.17 ± 0.02 |
| Copies, positivity, LTCH, weather, wastewater | 0.21 ± 0.04 | 0.17 ± 0.02 | 0.17 ± 0.02 |
| Copies, positivity, weather, wastewater, date | 0.19 ± 0.03 | 0.17 ± 0.02 | 0.18 ± 0.016 |
| Copies, positivity, LTCH, weather, wastewater, date | 0.21 ± 0.04 | 0.17 ± 0.02 | 0.18 ± 0.016 |
| **Area under the receiver operating curve (AUROC)** | | | |
| Floors: SC2 viral copies* | 0.82 ± 0.05 | NA | NA |
| Floors: SC2 positivity* | 0.81 ± 0.04 | NA | NA |
| Waste-water SC2:PMMoV 7-day endpoint average* | 0.82 ± 0.05 | NA | NA |
| Copies, positivity | 0.81 ± 0.04 | 0.73 ± 0.05 | 0.79 ± 0.04 |
| Copies, positivity, LTCH | 0.81 ± 0.05 | 0.79 ± 0.04 | 0.80 ± 0.04 |
| Copies, positivity, wastewater | 0.85 ± 0.04 | 0.83 ± 0.03 | 0.87 ± 0.03 |
| Copies, positivity, weather | 0.84 ± 0.04 | 0.83 ± 0.03 | 0.84 ± 0.03 |
| Weather covariates, wastewater | 0.81 ± 0.05 | 0.76 ± 0.08 | 0.79 ± 0.06 |
| Weather covariates | 0.74 ± 0.06 | 0.73 ± 0.07 | 0.74 ± 0.07 |
| Weather covariates, LTCH | 0.82 ± 0.05 | 0.75 ± 0.09 | 0.79 ± 0.05 |
| Copies, positivity, weather, wastewater | 0.87 ± 0.04 | 0.86 ± 0.03 | 0.87 ± 0.03 |
| Copies, positivity, LTCH, weather, wastewater | 0.88 ± 0.03 | 0.86 ± 0.03 | 0.87 ± 0.03 |
| Copies, positivity, weather, wastewater, date | 0.86 ± 0.04 | 0.86 ± 0.03 | 0.88 ± 0.03 |
| Copies, positivity, LTCH, weather, wastewater, date | 0.88 ± 0.03 | 0.86 ± 0.04 | 0.88 ± 0.03 |

Legend: AUROC = area under the receiver operating curve, SC2 = SARS-CoV-2, LTCH = long-term care home, PMMoV = pepper mild mottle virus. *Indicates a non-penalized logistic regression method was used for these single predictor models (using R's *glm* package), rather than LASSO. *LTCH* indicates the inclusion of a dummy variable for each site. *NA* indicates a combination that was not evaluated.

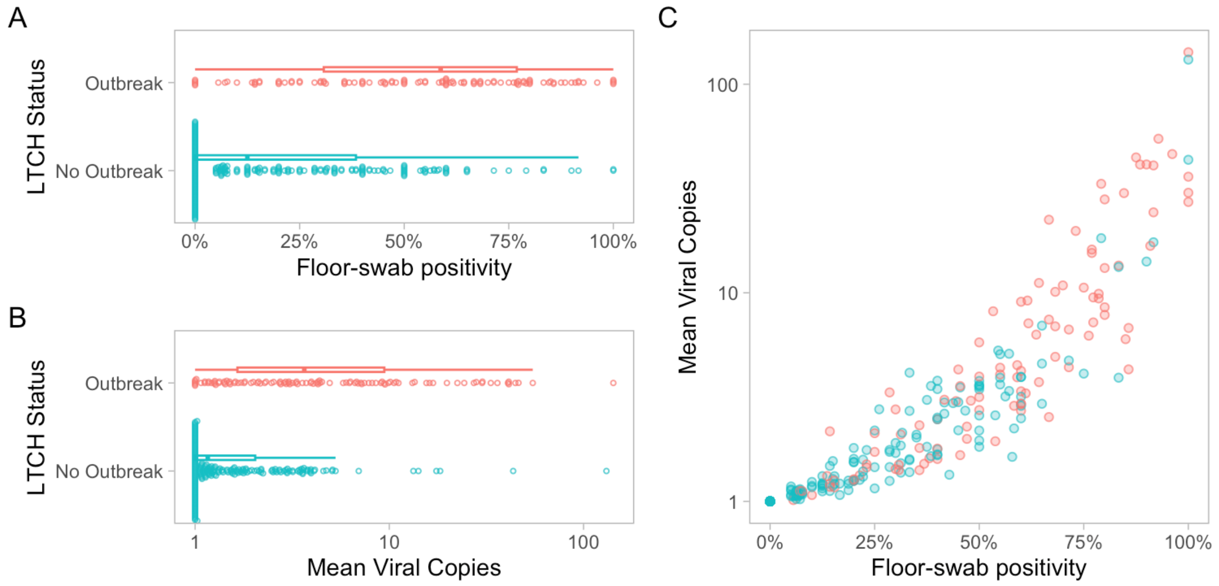


**Supplemental Figure S1**. Relationship between positivity and quantity of SARS-CoV-2 viral RNA detected on floor swabs collected at ten long-term care homes (LTCHs) by concurrent LTCH COVID-19 outbreak status. Red glyphs indicate the outbreak condition; blue indicates no outbreak.

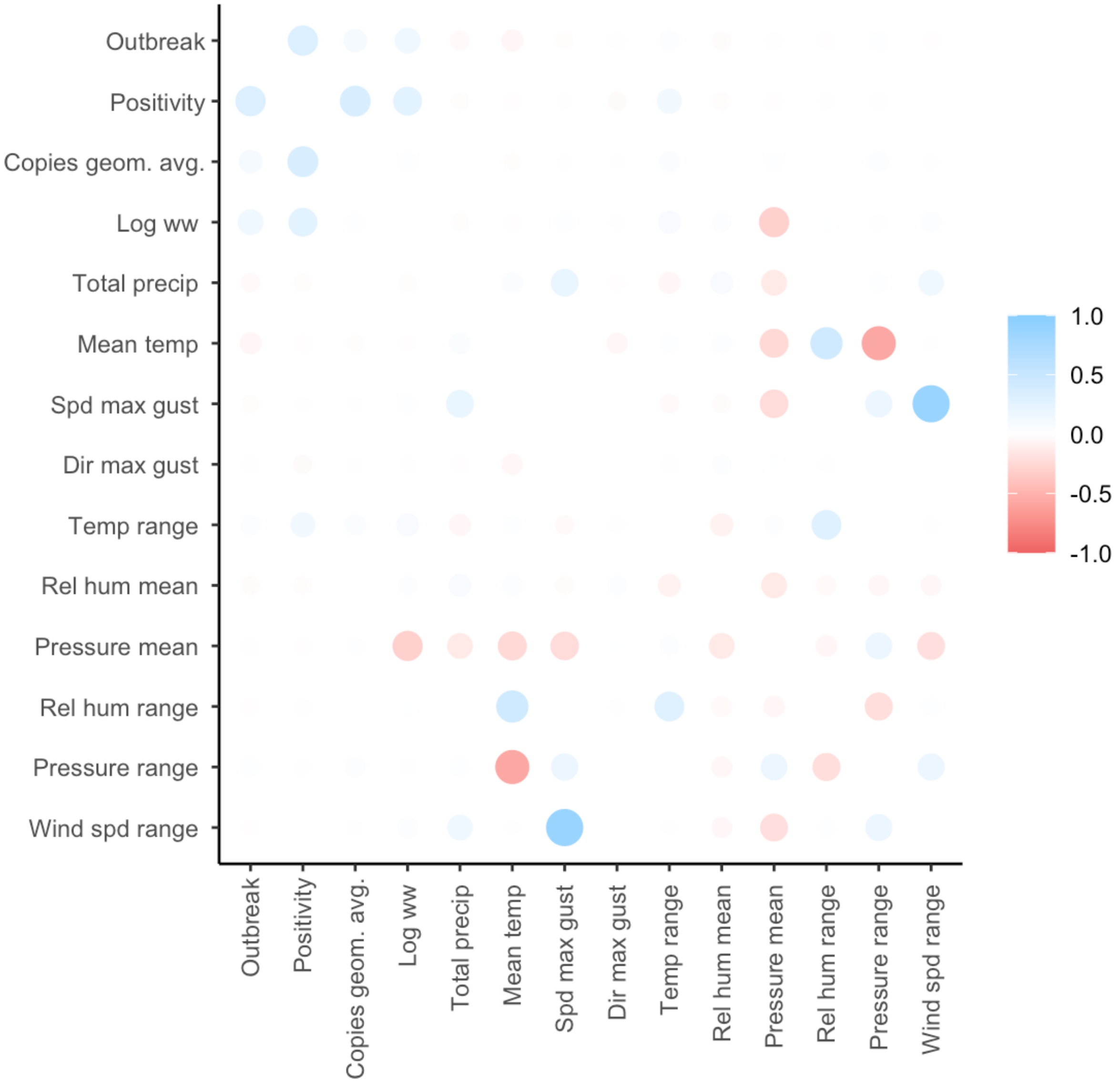


**Supplemental Figure S2**. Pearson correlations for long-term care home (LTCH) COVID-19 outbreak status and covariates from floor swabbing, building characteristics, regional wastewater, and regional weather. Copies geom. avg. = geometric mean number of SARS-CoV-2 RNA copies detected plus one. The pseudocount of one copy was added to each value of viral copies to enable logarithmic transformation of these data (as the logarithm of zero is undefined). Log ww = log transformed 7-day endpoint average wastewater.
